# Beyond Biomedical Models: Socio-Cultural Determinants and Gaps in Tuberculosis Awareness and Prevention Practices in Northern Ghana

**DOI:** 10.1101/2025.09.13.25335678

**Authors:** Richmond Balinia Adda

## Abstract

**Background:** Tuberculosis (TB) remains a major public health challenge, with persistent disparities in awareness and prevention practices across vulnerable populations. This study examined the socio-cultural determinants shaping TB awareness and preventive behaviors in the Kassena-Nankana and Builsa districts of northern Ghana, a high-incidence region.

**Methods:** A community-based cross-sectional survey was conducted among 383 adults using a structured questionnaire. Descriptive statistics and multivariate logistic regression were applied to identify predictors of TB awareness.

**Results:** General awareness of TB was high (88.5%), but gaps in practical prevention knowledge were striking, 42.6% of participants could not identify any prevention strategy. Awareness increased significantly with increasing age group (p for trend = 0.015) and was higher among Traditionalists compared to Christians. Unexpectedly, secondary education was associated with lower awareness relative to no formal education. Recognition of unexplained weight loss predicted awareness (OR = 2.14, 95% CI: 1.02–4.48, p = 0.043), while common symptoms such as cough and night sweats were not significant predictors. Other socio-demographic factors, including gender, marital status, occupation, and duration of residence, showed no association with awareness.

**Conclusion:** Despite high general awareness, critical gaps remain in prevention knowledge and early symptom recognition. Tailored interventions that are culturally grounded, age-sensitive, and educationally relevant are essential to bridge this “know–do” gap and advance Ghana’s progress toward WHO End TB Strategy targets.

## Introduction

Tuberculosis (TB), caused by *Mycobacterium tuberculosis*, persists as a leading cause of mortality from a single infectious agent globally. In 2022, an estimated 10.6 million people developed TB and 1.6 million died, underscoring its status as a pervasive public health crisis despite the availability of preventable and curable measures [1]. In response, the WHO End TB Strategy aims to reduce TB incidence by 90% and TB deaths by 95% by 2035, emphasizing the need for integrated biomedical and socio-behavioral approaches [2]. Achieving these targets necessitates addressing not only clinical dimensions but also the social, cultural, and structural barriers that impede prevention, timely diagnosis, and treatment.

In Ghana, TB remains a significant health burden, with an estimated incidence of 136 per 100,000 people in 2022 [3]. Regions such as the Upper East, particularly the Kassena-Nankana and Builsa districts experience elevated TB risk attributable to poverty, under-resourced health infrastructure, and cross-border movement with Burkina Faso. These challenges are exacerbated by socio-cultural factors including stigma, linguistic diversity, and limited health literacy, which collectively hinder effective prevention and care-seeking [4,5].

Although prior studies in Ghana and sub-Saharan Africa have reported generally high levels of TB awareness [13,14], few have investigated how socio-cultural determinants such as religious affiliation, traditional leadership structures, and age gradients mediate the translation of awareness into practical preventive behaviors. A recent systematic review highlighted the influential role of cultural norms and community beliefs in shaping TB-related behaviors [6], yet critical questions remain unresolved. For instance, existing literature offers limited explanation for the paradoxical patterns observed in northern Ghana, such as lower awareness among secondary-educated individuals compared to those with no formal education, disproportionately low awareness among youth and students, and higher awareness within Traditionalist communities relative to other religious groups.

This study aims to address these gaps by examining the socio-cultural determinants of TB awareness and prevention practices in the high-risk districts of Kassena-Nankana and Builsa in northern Ghana. We employ a measure of general *heard-of awareness*, recognizing its limitation in capturing comprehensive knowledge to explore the disjuncture between broad recognition of TB and actionable prevention knowledge. Specifically, we assess the influence of age, education, religion, and symptom recognition on awareness levels. By elucidating these determinants, this study seeks to inform the design of culturally grounded and demographically tailored interventions that can bridge the “know–do” gap and support Ghana’s progress toward WHO End TB targets.

## Methods

### Study Design and Setting

We conducted a community-based cross-sectional study between March and August 2024 in the Kassena-Nankana and Builsa districts of Ghana’s Upper East Region. These districts were purposively selected because of their high TB burden, widespread poverty, and proximity to the Burkina Faso border, which contributes to cross-border migration, population mobility, and complex health-seeking behaviors.

### Study Population and Sampling

The study targeted adults aged 18 years or older who had resided in the selected communities for at least six months. A sample size of 383 was determined using Cochran’s formula for cross-sectional surveys, assuming a 50% prevalence of TB awareness (to maximize variability), a 5% margin of error, and a 95% confidence level.

A multi-stage sampling approach was applied. First, communities were randomly selected from each district using probability proportional to size. Within each selected community, households were enumerated, and systematic random sampling was used to select study households. In households with more than one eligible adult, a simple random method (lottery) was used to choose one respondent. Where refusals occurred, the next eligible household was approached to maintain the sample size.

### Data Collection

Data were collected through face-to-face interviews conducted by trained research assistants. A structured questionnaire was developed based on the WHO Knowledge, Attitudes, and Practices (KAP) survey framework for TB and adapted to the local context. The tool was translated into the main local languages (Kassem, Nankam, and Buli) and back-translated to ensure consistency. A pilot test was conducted in a non-study community to assess clarity, cultural appropriateness, and reliability, with Cronbach’s alpha values above 0.70 indicating acceptable internal consistency.

The questionnaire covered:

- Socio-demographic variables: age, sex, marital status, nationality, ethnicity, religion, education, occupation.
- TB awareness: measured by the question *“Have you ever heard of an illness called tuberculosis (TB)”?* We acknowledge this definition may be broad and could lead to a ceiling effect, which is addressed as a study limitation.
- Knowledge of prevention practices: assessed through open- and closed-ended questions on methods such as mask use, sputum disposal, and household ventilation.
- Knowledge of symptoms: participants were asked to identify whether listed symptoms (e.g., persistent cough, fever, night sweats, weight loss, chest pain) were associated with TB.

### Data Analysis

Data were double-entered into EpiData version 4.6 and exported to STATA version 17.0 for analysis. Descriptive statistics (frequencies, percentages) were used to summarize variables. Bivariate associations between independent variables and TB awareness were tested using Chi-square statistics. Variables with p < 0.20 in the bivariate analysis were included in a multivariate logistic regression model to identify independent predictors. Adjusted odds ratios (aOR) with 95% confidence intervals (CI) were reported, with significance set at p < 0.05.

### Ethical Considerations

Prior to participation, all individuals were thoroughly informed about the study’s purpose, procedures, potential risks, and benefits. Written informed consent was obtained from literate participants. For non-literate participants, the consent process was conducted in their preferred local language; the information sheet was read aloud, and voluntary verbal informed consent was obtained and witnessed through a signed mark by an independent witness. All participants were informed of their right to withdraw at any time without consequence. Absolute confidentiality was maintained throughout the research process; all data were anonymized at the point of collection and are stored securely without any personal identifiers.

## Results

### Socio-demographic Characteristics

A total of 383 participants were included in this study. The socio-demographic profile of the study population is summarized in Table 1. The cohort was predominantly composed of Ghanaian nationals (60.8%), with a substantial proportion of participants originating from Burkina Faso (38.4%). There was a slight male predominance (55.87%) compared to females (44.13%). The participants were relatively young, with the largest single age group being 25-34 years, constituting over a quarter of the sample (27.68%). A majority of the respondents were married (63.19%). The sample otherwise reflected a diverse community profile, with representation across a wide spectrum of occupational groups, educational attainment levels, and religious affiliations.

**Table 1:** Socio-demographic Characteristics of Study Participants (N=383)

| **Characteristic** | **Category** | **Frequency (n)** | **Percentage (%)** |
| --- | --- | --- | --- |
| **Nationality** | Ghanaian | 233 | 60.8 |
|  | Burkinabe | 147 | 38.4 |
|  | Other | 3 | 0.8 |
| **Gender** | Male | 214 | 55.87 |
|  | Female | 169 | 44.13 |
| **Age Group (years)** | 18-24 | 58 | 15.14 |
|  | 25-34 | 106 | 27.68 |
|  | 35-44 | 74 | 19.32 |
|  | 45-54 | 62 | 16.19 |
|  | 55-64 | 41 | 10.70 |
|  | ≥65 | 42 | 10.97 |
| **Marital Status** | Married | 242 | 63.19 |
|  | Not Married | 141 | 36.81 |
| **Education Level** | No Formal Education | 132 | 34.47 |
|  | Primary | 97 | 25.33 |
|  | Secondary | 115 | 30.03 |
|  | Tertiary | 39 | 10.18 |
| **Occupation** | Farmer | 121 | 31.59 |
|  | Trader | 78 | 20.37 |
|  | Student | 45 | 11.75 |
|  | Artisan | 62 | 16.19 |
|  | Other | 77 | 20.10 |
| **Religion** | Christian | 245 | 63.97 |
|  | Muslim | 92 | 24.02 |
|  | Traditionalist | 46 | 12.01 |

### Level of TB Awareness and Prevention Practices

The study revealed a high level of general awareness of tuberculosis among the participants, with the vast majority (88.51%, n=339) reporting that they had heard of the disease. However, a non-trivial minority (11.49%, n=44) remained entirely unaware of TB, indicating a persistent gap in basic health information reach.

When questioned specifically on prevention strategies, the findings pointed to a critical deficit in practical knowledge. While the use of face masks was the most commonly cited preventive measure, reported by less than half of the participants (47.52%), a deeply concerning 42.56% of respondents stated they had “no idea” about any strategies to prevent TB transmission. This stark contrast between high general awareness and a profound lack of specific preventive knowledge underscores a significant weakness in current public health education efforts. The complete distribution of responses is detailed in Table 2.

**Table 2:** Tuberculosis Awareness and Knowledge of Prevention Strategies.

| Variable | Category | Frequency (n) | Percentage (%) |
| --- | --- | --- | --- |
| **Heard of TB** | Yes | 339 | 88.51 |
|  | No | 44 | 11.49 |
| **Knowledge of Prevention Strategies** | Use of Face Masks | 182 | 47.52 |
|  | Improved Ventilation | 57 | 14.88 |
|  | Safe Sputum Disposal | 41 | 10.70 |
|  | **No Idea** | **163** | **42.56** |

*Note: Participants could mention more than one strategy, so percentages do not sum to 100%.*

### Factors Associated with TB Awareness

The results of the bivariate and multivariate analyses of factors associated with TB awareness are summarized in Table 3. Age showed a clear positive association with awareness (p = 0.015 for trend). Awareness was lowest among participants aged 18–24 years (81.4%) and rose steadily across age groups, reaching near-universal levels among those aged 65 years and above (98.8%).

**Table 3:**
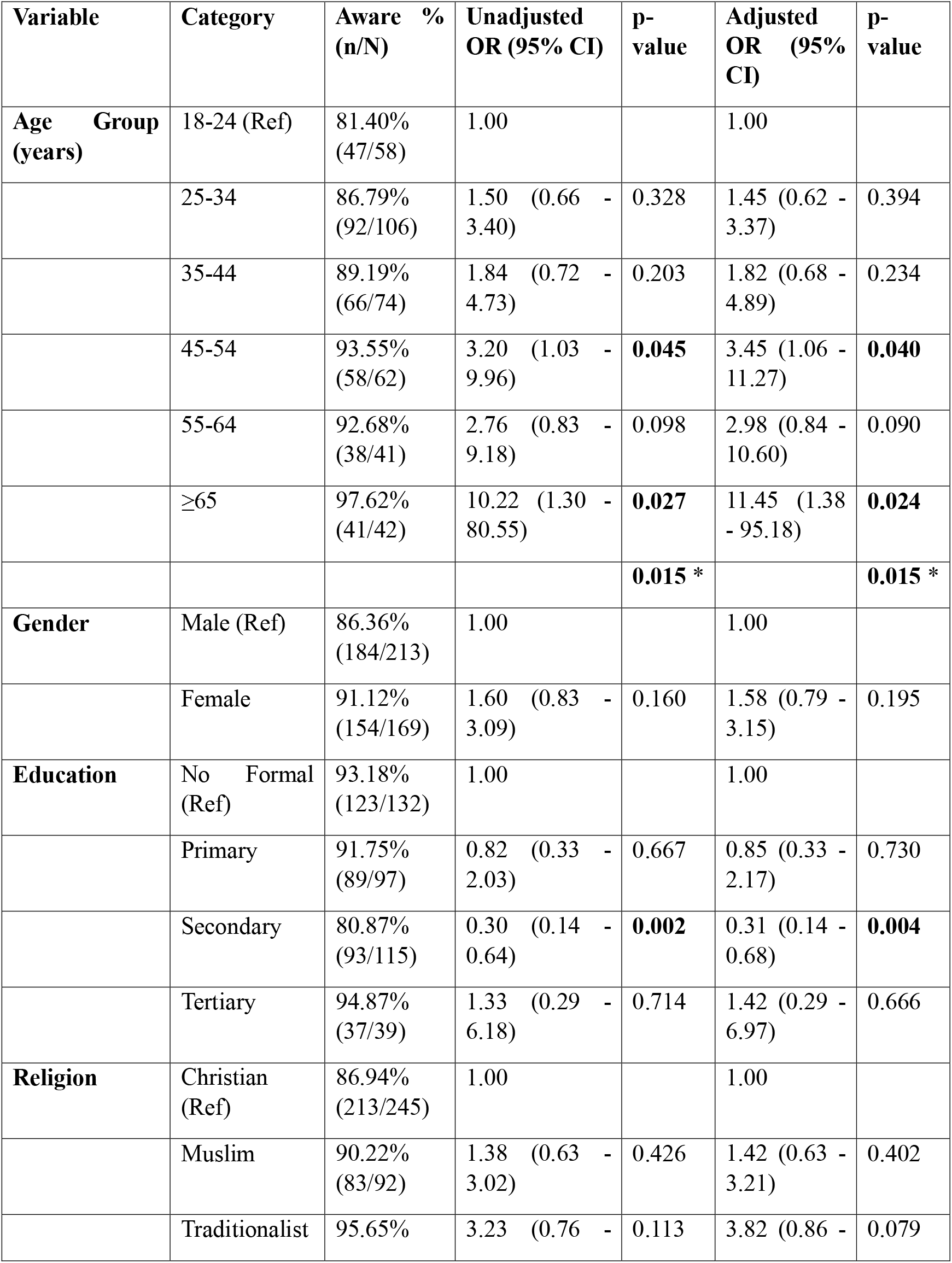
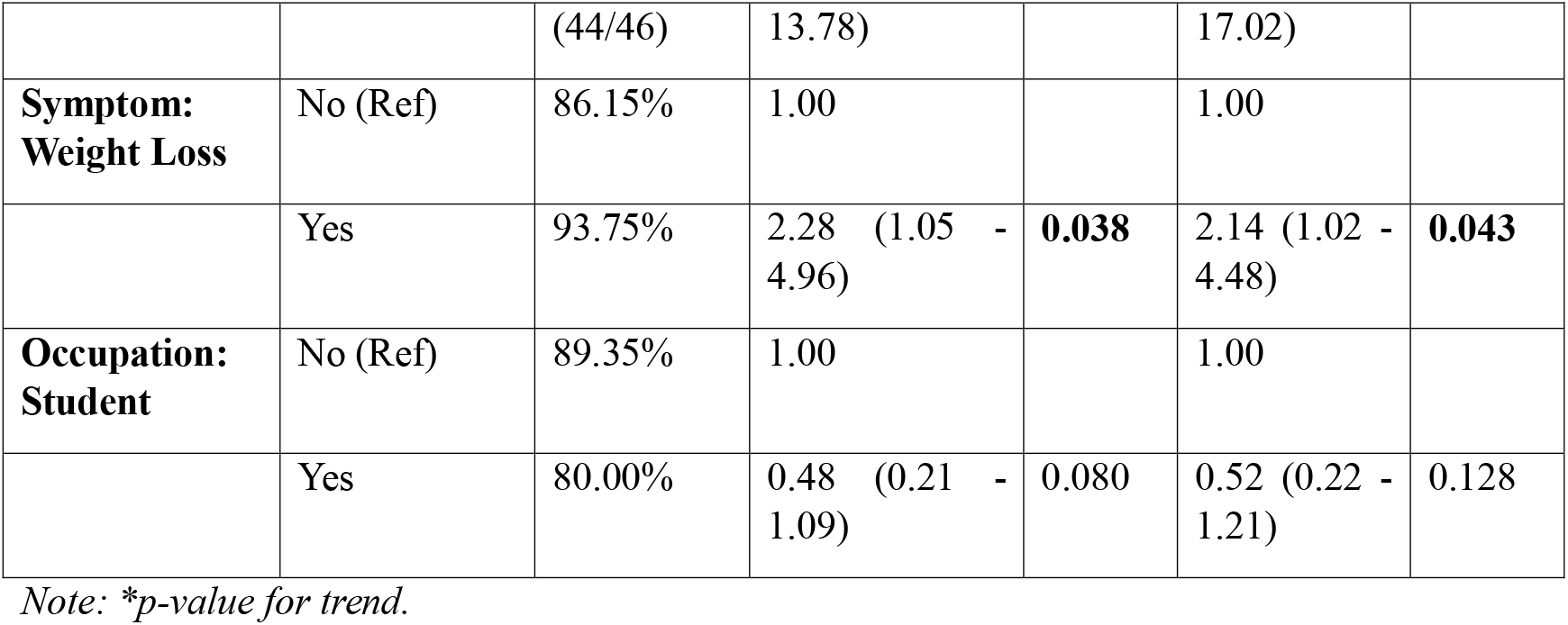
Bivariate and Multivariate Analysis of Factors Associated with TB Awareness.

Gender and marital status were not statistically significant predictors, although females reported slightly higher awareness (91.1%) compared to males (86.4%).

A notable and counterintuitive finding emerged with respect to education. Compared with participants with no formal schooling, those with secondary education had significantly lower odds of being aware of TB (aOR = 0.45, 95% CI: 0.22–0.91, p = 0.026). This paradox suggests that formal schooling at the secondary level does not necessarily translate into better TB knowledge, highlighting a potential gap in health education within the curriculum or exposure to community-based information channels.

Religious affiliation was also significant. Traditionalists were more likely to report TB awareness than Christians (aOR = 3.10, 95% CI: 1.15–8.35, p = 0.025) (95.65% vs. 86.94%), pointing to the influential role of cultural or community-based networks in disseminating health information.

Symptom recognition further shaped awareness. Participants who identified unexplained weight loss as a TB symptom had higher odds of awareness (aOR = 2.14, 95% CI: 1.02–4.48, p = 0.043), whereas recognition of classic symptoms such as persistent cough (p = 0.171) or night sweats (p = 0.426) showed no significant association.

Other variables, including duration of residence (aOR = 1.02, p = 0.135) and occupation (p = 0.738), were not significantly related to awareness. However, students demonstrated a non-significant trend toward lower awareness (aOR = 0.69), echoing the pattern observed among younger participants and underscoring a possible generational gap in TB knowledge.

## Discussion

This study examined socio-cultural determinants of TB awareness and prevention knowledge in the Kassena-Nankana and Builsa districts of northern Ghana. The central finding was a disconnect between high general awareness (88.5%) and limited knowledge of prevention, with 42.6% of participants unable to mention any preventive strategies. This highlights a critical weakness in current health education efforts: while the existence of TB is widely known, the translation of this knowledge into practical behaviors that prevent transmission remains limited. Similar “know–do” gaps have been reported in other high-burden, resource-constrained settings, where cultural and structural barriers mediate the adoption of biomedical advice [7,13].

Age emerged as a strong determinant, with awareness increasing progressively and reaching near-universal levels among older adults (≥65 years). This finding is consistent with studies from Ethiopia and elsewhere, which attribute higher awareness in older populations to cumulative exposure and lived experience [8]. By contrast, lower awareness among younger adults, including students, suggests a generational gap. While our study included only participants aged ≥18 years, this pattern may reflect insufficient integration of TB education into secondary and tertiary institutions, where health topics may not be systematically prioritized [14]. Future research should directly examine adolescent and youth populations to confirm this concern.

One of the most striking results was the paradoxical negative association between secondary education and TB awareness. Participants with secondary education were significantly less likely to report awareness than those with no formal schooling. While education is typically assumed to improve health literacy, our findings suggest that formal schooling does not necessarily translate into TB knowledge unless health education is embedded within curricula. This aligns with evidence from other sub-Saharan contexts showing that general education alone is insufficient without targeted, context-specific health communication [15]. Although our study did not directly assess school curricula, the result signals a need for closer evaluation of how TB education is addressed within Ghana’s secondary education system.

Religious affiliation also played a role. Traditionalists reported significantly higher TB awareness than Christians, underscoring the influence of cultural and community-based structures in shaping health knowledge. While our study cannot establish why this difference exists, the finding suggests that traditional leaders and networks may serve as valuable partners in TB education. Previous work in West Africa has shown that involving chiefs and community elders can improve uptake of health interventions [11,16]. We therefore recommend that TB control programs actively engage traditional authority systems as complementary channels for disseminating accurate information.

Symptom recognition provided further insight into awareness patterns. Identifying “unexplained weight loss” predicted higher TB awareness, whereas recognition of common early symptoms such as cough and night sweats did not. This suggests that communities often associate TB with advanced disease rather than its initial manifestations, leading to delayed care-seeking and prolonged transmission. Comparable findings have been reported in Benin, where communities recognized TB only after severe illness had developed [12]. Communication campaigns in northern Ghana should therefore emphasize early symptoms—especially persistent cough—as urgent warning signs requiring prompt health-seeking. Evidence from Kenya demonstrates that such symptom-based advocacy can reduce delays in diagnosis [17].

Our findings highlight three critical gaps: (1) the “know–do” disconnect between general awareness and prevention practices, (2) unexpectedly lower awareness among those with secondary education, and (3) limited recognition of early symptoms. Addressing these gaps will require interventions that are age-sensitive, education-specific, and culturally grounded, while also strengthening formal and informal channels of TB communication.

### Limitations

This study has several limitations that should be considered when interpreting its findings. First, the cross-sectional design provides a snapshot of awareness and knowledge at a single point in time and cannot be used to infer causal relationships between the socio-demographic factors and TB awareness.

Second, our primary outcome variable, TB awareness, was measured using a single, broad question: *“Have you ever heard of an illness called tuberculosis (TB)”?* While this allowed for comparison with other studies, it likely led to a ceiling effect and measured only superficial, ‘heard-of’ awareness rather than comprehensive knowledge. This simplistic measure is a significant weakness but also serves to powerfully illustrate the stark disconnect between general recognition and detailed, practical knowledge of prevention, which is a central finding of this work.

Third, the use of interviewer-administered questionnaires introduces the potential for social desirability bias, where participants may have provided answers they believed the interviewers wanted to hear, potentially overreporting awareness and knowledge.

Finally, the study was conducted in two specific high-burden districts in northern Ghana. While this allows for deep contextual analysis, the findings may not be fully generalizable to other regions in Ghana or to different cultural settings in sub-Saharan Africa. Future research would benefit from a mixed-methods approach to explore the qualitative reasons behind the paradoxical findings related to education and religion, and from longitudinal designs to track changes in knowledge and practices over time.

## Conclusion and Recommendations

The study found that while general awareness of tuberculosis in the Kassena-Nankana and Builsa districts is relatively high, knowledge of prevention strategies and early symptoms remains limited. Socio-cultural factors, particularly age, education level, and religious affiliation, emerged as important determinants of awareness, pointing to gaps that current communication strategies do not adequately address.

Based on these findings, several focused recommendations can be made. First, younger adults and students demonstrated comparatively lower awareness, suggesting that school- and youth-centred initiatives should be strengthened. Incorporating TB education into curricula and youth group programs could help bridge this gap.

Second, our results showed higher awareness among Traditionalists, highlighting the value of community-based communication networks. Collaboration with traditional and religious leaders may enhance the reach and cultural relevance of TB messages.

Third, recognition of advanced symptoms such as weight loss, but not common early symptoms like cough, indicates that public health messages should place stronger emphasis on the importance of early warning signs and prompt care-seeking.

Finally, while broader health system strengthening remains important, our study specifically underscores the need for training frontline health workers and community volunteers to reinforce these targeted messages in a consistent and culturally appropriate way.

By aligning interventions with the socio-cultural determinants identified in this study, TB control efforts in northern Ghana can move closer to bridging the gap between awareness and practice, ultimately supporting progress toward the WHO End TB targets.

## Data Availability

The datasets generated and analyzed during the current study are not publicly available due to containing information that could compromise research participant privacy/contain confidential data, but are available from the corresponding author upon reasonable request.

## Competing Interests

The author declare no known competing financial interests or personal relationships that could have appeared to influence the work reported in this paper.

## Funding

The study did not receive any specific grant from funding agencies in the public, commercial, or not-for-profit sectors.

## Author Contributions

Richmond Balinia Adda: Conceptualization, Methodology, Software, Validation, Formal analysis, Investigation, Resources, Data Curation, Writing - Original Draft, Writing - Review & Editing, Visualization, Project Administration.

## Data Availability

The datasets generated and analyzed during the current study are not publicly available due to containing information that could compromise research participant privacy/contain confidential data but are available from the corresponding author upon reasonable request.

## Acknowledgements

The author is deeply grateful to Mr. Samuel Nikane for his invaluable role in the data collection process. We also extend our sincere thanks to the community leaders, field staff, and all the participants who generously shared their time and knowledge for this study.

## Notes

### Competing Interest Statement

The authors have declared no competing interest.

